# Expanding The Use Of Friedewald Equation In Lipid Profile Testing Among HIV Positive And Negative Elderly Persons

**DOI:** 10.1101/2021.09.08.21263022

**Authors:** Emunyu Jude, Semigga Brian, Kisembo Stephen, Namukwaya Brenda, Namubiru Bridget, Bakyayita Charles, Emmanuel Omony

## Abstract

Hypercholesterolemia and hypocholesterolemia are associated with mortality which warrants routine lipid profile testing. This financially burdens the already overwhelmed health sector especially in developing countries. Additionally, lipid profile test reagent stock-out or failure to afford all tests affects result interpretation. In 1972, James Friedewald published a statistical model to calculate low density lipo-protein. The study aim was to determine the percentage error of the James Friedewald equation in calculating all lipid profile test parameters. A retrospective study from 2018 was performed at Mildmay Uganda involving lipid profile results of 103 persons (48 HIV-positive and 55 HIV-negative) 50 years and older enrolled in a previous cross-sectional study. The Friedewald equation was used to calculate total cholesterol, high density lipoprotein, triglycerides and low density lipo-protein. The percentage error of calculated values in reference to measured values was ascertained. Pearson correlation between measured and calculated results was determined among all persons and classified by HIV status. The total error of calculated analytes was 7% (low density lipo-protein), 17% (high density lipo-protein), 39% (triglycerides) and 4% (total cholesterol). Pearson correlations were 0.98 (all persons), 0.98 (HIV-negative) and 0.98 (HIV-positive) for low density lipo-protein, 0.89 (all persons), 0.90 (HIV-positive) and 0.88 (HIV-negative) for high density lipo-protein, 0.75 (all persons), 0.76 (HIV-negative) and 0.77 (HIV-positive) for triglycerides, 0.99 (all persons), 0.98 (HIV-negative) and 0.99 (HIV-positive) for total cholesterol. In conclusion, Friedewald equation reliably calculated low density lipo-protein, total cholesterol (most accurate) and high density lipo-protein while triglycerides calculation was erroneous among persons aged ≥ 50 years.

## 1.0 INTRODUCTION

Cardiovascular diseases are estimated to cause 17.3 million deaths globally per year and are predicted to account for over 23.6 million global deaths per year by 2030 ^1^. As much as the globe remains poised with a threat of mortality from cardiovascular disease, three quarters of these deaths occur in low and middle-income countries ^2,3^. Hypercholesterolemia has been established as an independent risk marker for the development of cardiovascular disease ^4^. However, hypercholesterolemia is affected by factors such as old age and medications such as anti-retroviral therapy among HIV positive persons. ^4,5^. Contrary to the link between hypercholesterolemia and cardiovascular disease, an association between hypocholesterolemia with a higher risk of mortality has also been demonstrated ^6^. This concept was as well supported by a study conducted in Finland that highlights low blood cholesterol levels to be linked with a lower survival rate than high blood cholesterol ^7^. These facts continue to support the need for routine lipid profile testing to monitor cholesterol levels. Clinical lipid profile testing entails serum analysis for total cholesterol, high density lipo-protein cholesterol (HDL-C), low density lipo-protein cholesterol (LDL-C) and triglycerides levels ^8^. High LDL-C has been demonstrated as an independent risk factor for development of cardiovascular disease making its quantification vital ^9^. Furthermore, reduction in blood LDL-C levels has been established as a good prognostic marker for statin therapy in management of hypercholesterolemia ^9,10^. Unlike other lipid profile tests that are only quantified by analytical methods, LDL-C can as well be calculated from a statistical model designed by James Friedewald in 1972 which continues to be widely used in clinical set ups ^11^. Low HDL-C on the other hand remains a powerful predictor of coronary heart disease residual risk even at low LDL-C levels and in conjunction with elevated triglycerides are considered high risk factors for coronary heart disease despite low LDL-C levels ^12^. A wholistic approach entailing analysis of all lipid profile parameters is thus essential. This however possess an increased economic burden to the already overwhelmed health sector especially in the developing countries. Moreover, laboratories in the developing countries are persistently faced with reagent stock outs and in some cases may not be able to perform all the four lipid profile tests. The Friedewald equation is used to calculate LDL-C using measured results of the other three lipid profile parameters; HDL-C, triglycerides and total cholesterol. This study re-arranged the Friedewald equation so as to be able to calculate any of the four lipid profile parameters when three of the other parameters have been measured hence expanding its utility.

## 2.0 MATERIALS AND METHODS

### 2.1 Study design and site

This was a retrospective study from 2018 which expanded the use of Friedewald equation from only LDL-C calculation to include calculation of other lipid profile parameters; total cholesterol, triglycerides and HDL-C. This study was performed at Mildmay Uganda. The study utilized data from the Neurocognitive decline and non-communicable chronic disease in older HIV infected adults on cART and HIV negative age/sex matched community controls-A pilot investigation performed at Mildmay Uganda in 2017 and 2018 (not published). A total number of 200 participants were enrolled in the study from which the data was extracted with inclusion criteria entailing having a non-detectable viral load for HIV positive individuals while both HIV negative and positive individuals were supposed to be 50 years and older, have consented and have no evidence of metabolic conditions. Exclusion criteria were detectable viral loads, being younger than 50 years, not consenting and having evidence of metabolic conditions.

### 2.2 Sample size

We conducted a census of all available data from the primary study. Data was analyzed from 103 lipid profile results that included 55 HIV negative and 48 HIV positive individuals, 35 males and 68 females with a minimum age of 50 years.

### 2.3 Inclusion and exclusion criteria

Inclusion into the study required results to have all the four lipid profile parameters (total cholesterol, triglycerides, HDL-C and LDL-C) measured and triglyceride levels below 4.52 mmol/L (400mg/dl) as this is a widely used triglyceride cut-off point to apply the Friedewald equation ^11^. Results with triglyceride values greater than 4.52 mmol/L (400mg/dl) were excluded from the study. Results that did not have all the 4 parameters i.e. HDL-C, LDL-C, triglycerides and total cholesterol measured were also excluded.

### 2.4 Sample collection, biochemical assays

After an 8 to 12 hour overnight fast by participants, a volume of 4mls of venous blood was taken into plain tubes by phlebotomy. The blood was then centrifuged at 3000 rpm for 5 minutes to obtain serum which was analyzed using the Roche Cobas C311 chemistry analyzer. Quantification was performed using enzymatic methods i.e. CHOD/PAP for total cholesterol, PEG/CHOD/PAP for HDL-C test, GEP/PAP for triglycerides and homogeneous test for LDL-C.

### 2.5 Expanding Friedewald equation to calculate LDL-C, HDL-C, triglyceride and total cholesterol

Friedewald equation ^11^ was used to calculate LDL-C, total cholesterol, HDL-C and triglycerides concentrations in mmol/L using three measured parameters as follows;

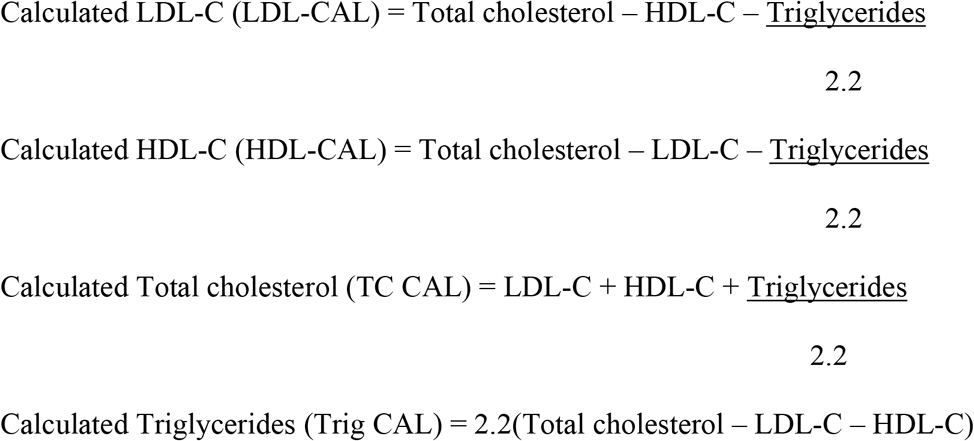

### 2.6 Establishing total error of calculated test results

Total error (TE) of Friedewald equation calculated HDL-C, LDL-C, total cholesterol and triglycerides was determined and compared to published total allowable error (TAE) of these analytes using the following equation ^13^.

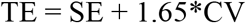

where; SE is the bias/systemic error, CV is imprecision from similar sample repeated analysis The mean systemic error of calculated LDL-C, HDL-C, total cholesterol and triglycerides using Friedewald equation was determined using measured results as reference from equation^14^:

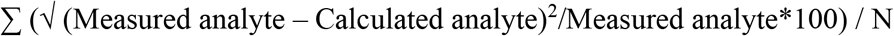

where; N= number of participant results used i.e. 103

Unlike analysis of a same sample that yields different results, calculating yields the same result as long as the measured values are un altered and thus the SE (bias/systemic error) was used as the TE in this case.

### 2.7 Correlation between measured and calculated values

Correlations between the measured and calculated values from the Friedewald equation for HDL-C, triglycerides, LDL-C and total cholesterol were ascertained. The correlation between these values classified based on HIV status was also established. Average correlations between measured and calculated lipid profile tests for HIV positive and HIV negative individuals were compared using the t-test. Correlations were established by statistical analysis using STATA software version 12 for windows.

### 2.8 Accuracy of expanded Friedewald equation in cardiovascular disease risk stratification

The sensitivity, specificity and likelihood ratios of Friedewald equation in calculating LDL-C, HDL-C and total cholesterol across cardiovascular risk assessment concentrations of these analytes stated in the third report of the National Cholesterol Education Program expert panel on detection evaluation and treatment of high blood cholesterol in Adults: Adult treatment panel III (NCEP ATP III guidelines) ^15^ were ascertained and from these the percentage of correctly classified subjects by Friedewald equation applied to the different calculated test results was determined.

### 2.9 Data analysis

MS Excel spread sheet was used to store data, calculate HDL-C, LDL-C, total cholesterol and triglyceride concentrations using the Friedewald equation and calculate the systemic error between measured and calculated values. Using STATA software version 12 for windows, Pearson’s correlation analysis was used to determine the correlation between measured and calculated lipid profile values for all participants and participants classified based on HIV status. For all statistical comparisons, a p < 0.05 was considered as statistically significant. Receiver operating characteristic (ROC) STATA analysis to determine sensitivity, specificity and likelihood ratios of Friedewald equation in calculating LDL-C, HDL-C and total cholesterol across the NCEP ATP III established concentrations for cardiovascular disease risk stratification. The LDL-C, HDL-C and total cholesterol concentrations in these categories were used as cut-offs.

### 2.10 Ethical consideration

This study was approved by the Mildmay Uganda research ethics committee. The study from which data was extracted was approved by Mildmay Uganda research ethics committee and carried out in accordance with the code of ethics of the world medical association (Declaration of Helsinki).

## 3.0 RESULTS

### 3.1 Baseline characteristics of study population

The population studied was a total of 103 participant results who had a minimum age of 50 years, 48 HIV positive and on ART, 55 HIV negative, 35 males and 68 females. The mean results were 5.07 mmol/L TC-M, 5.22 mmol/L TC-CAL, 1.44 mmol/L HDL-M, 1.29 mmol/L HDL-CAL, 1.35 mmol/L Trig-M, 1.01 mmol/L Trig-CAL, 3.17 mmol/L LDL-M and 3.01 mmol/L LDL-CAL while the standard deviations were 1.13 mmol/L TC-M, 1.19 mmol/L TC-CAL, 0.43 mmol/L HDL-M, 0.46 mmol/L HDL-CAL, 0.57 mmol/L Trig-M, 0.69 mmol/L Trig-CAL, 1.07 mmol/L LDL-M and 1.02 mmol/L LDL-CAL as shown in table 1. P-values from comparison of means by unpaired t-test yielded significant values of 0.3408 between TC-M and TC-CAL, 0.2880 between LDL-M and LDL-CAL.

**Table 1:** Mean, standard deviation, maximum and minimum values of measured test results, calculated test results and total errors of calculated lipid profile test results

| **VARIABLE** | **MEAN** | **SD** | **MEDIAN** | **MAX** | **MIN** |
| --- | --- | --- | --- | --- | --- |
| **TC_M** | 5.06767 | 1.132134 | 5.09 | 9.12 | 2.09 |
| **TC_CAL** | 5.222427 | 1.193355 | 5.09 | 9.13 | 2.145455 |
| **TC_TOTAL ERROR** | 4.333832 | 2.346128 | 4.347826 | 10.76299 | 0 |
| **HDL_M** | 1.44233 | .4321474 | 1.34 | 2.8 | .65 |
| **HDL_CAL** | 1.287573 | .4560769 | 1.220909 | 2.929091 | .5227273 |
| **HDL_TOTAL ERROR** | 16.22477 | 10.21387 | 15.74508 | 53.74095 | 0 |
| **TRIG_M** | 1.349903 | .573733 | 1.24 | 3.22 | .61 |
| **TRIG_CAL** | 1.009437 | .6932297 | .836 | 3.058 | -.176 |
| **TRIG_TOTAL ERROR** | 39.98185 | 26.41784 | 36.47887 | 123.1579 | 2.88e-14 |
| **LDL_M** | 3.166505 | 1.067985 | 3.1 | 6.79 | .68 |
| **LDL_CAL** | 3.011748 | 1.01623 | 3.030909 | 6.78 | .6863636 |
| **LDL_TOTAL ERROR** | 7.149492 | 3.933574 | 6.911693 | 18.18182 | 0 |

Key: LDL-M; measured LDL-C, LDL-CAL; calculated LDL-C, Trig-M; measured triglycerides, Trig-CAL; calculated triglycerides, HDL-M; measured HDL-C, HDL-CAL; calculated HDL-C, TC-M; measured total cholesterol, TC-M; measured total cholesterol

### 3.2 ESTIMATION OF TOTAL ERROR

The mean total error of LDL-CAL, HDL-CAL, Trig-CAL and TC-CAL in comparison to their directly quantified values was ±7.15%, ±16.22%, ±39.98% and ±4.33% respectively (table1).

### 3.3 CORRELATION BETWEEN MEASURED AND CALCULATED RESULTS

Pearson correlation between TC-CAL and TC-M was 0.9852, correlation between HDL-CAL and HDL-M was 0.8911, between Trig-CAL and Trig-M was 0.7532 while that between HDL-CAL and HDL-M was 0.9811 (table 2). Scatter plots of measured against calculated values are shown in figures 1. HIV positive persons had an average correlation of 0.906825 which was statistically similar to the average correlation 0.90505 seen for HIV negative persons with a p-value of 0.9877. TC-CAL and TC-M showed the highest correlation in both groups i.e. 0.9810 and 0.9901 among HIV negative and positive individuals respectively while trig-M and trig-CAL showed the lowest correlations i.e. 0.7601 and 0.7785 among HIV negatives and positives respectively (table 3). Pearson correlations were 0.9794 (HIV negative) and 0.9847 (HIV positive) for low density lipo-protein while for high density lipo-protein 0.9021 (HIV positive) and 0.8810 (HIV positive).

**Table 2:** Correlation between measured and calculated lipid profile results (significance in italics)

| **ANALYTES** | **TC_M** | **HDL_M** | **TRIG_M** | **LDL_M** |
| --- | --- | --- | --- | --- |
| **TC-CAL** | 0.9853* |  |  |  |
|  | *0.0000* |  |  |  |
| **HDL-CAL** |  | 0.8911* |  |  |
|  |  | *0.0000* |  |  |
| **TRIG CAL** |  |  | 0.7532* |  |
|  |  |  | *0.0000* |  |
| **LDL-CAL** |  |  |  | 0.9812* |
|  |  |  |  | *0.0000* |

**Key:** LDL-M; measured LDL-C, LDL-CAL; calculated LDL-C, Trig-M; measured triglycerides, Trig-CAL; calculated triglycerides, HDL-M; measured HDL-C, HDL-CAL; calculated HDL-C, TC-M; measured total cholesterol, TC-M; measured total cholesterol

**Table 3:**
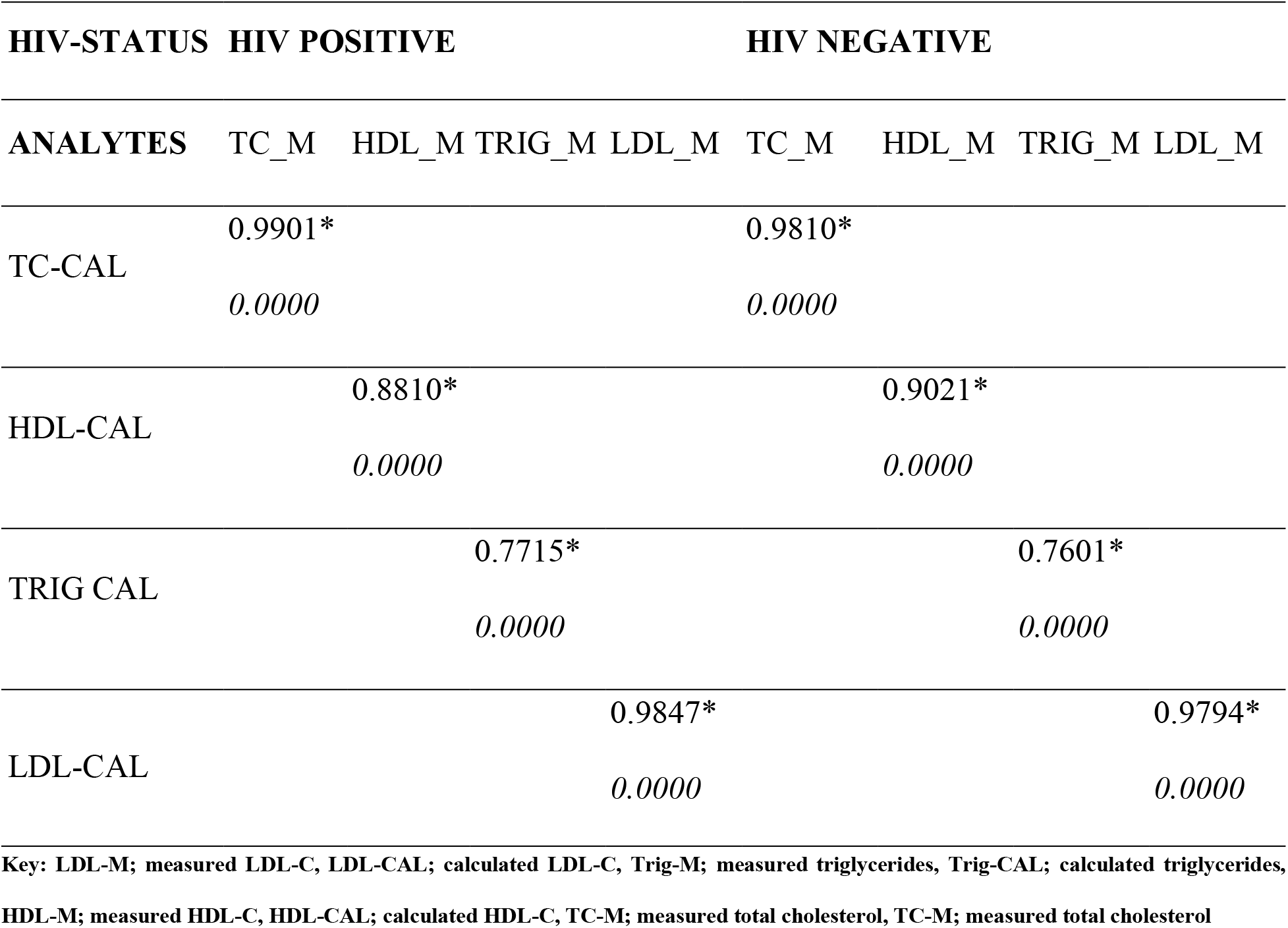
Correlation between measured and calculated lipid profile results across different HIV status (significance in italics)

**Figure1:**
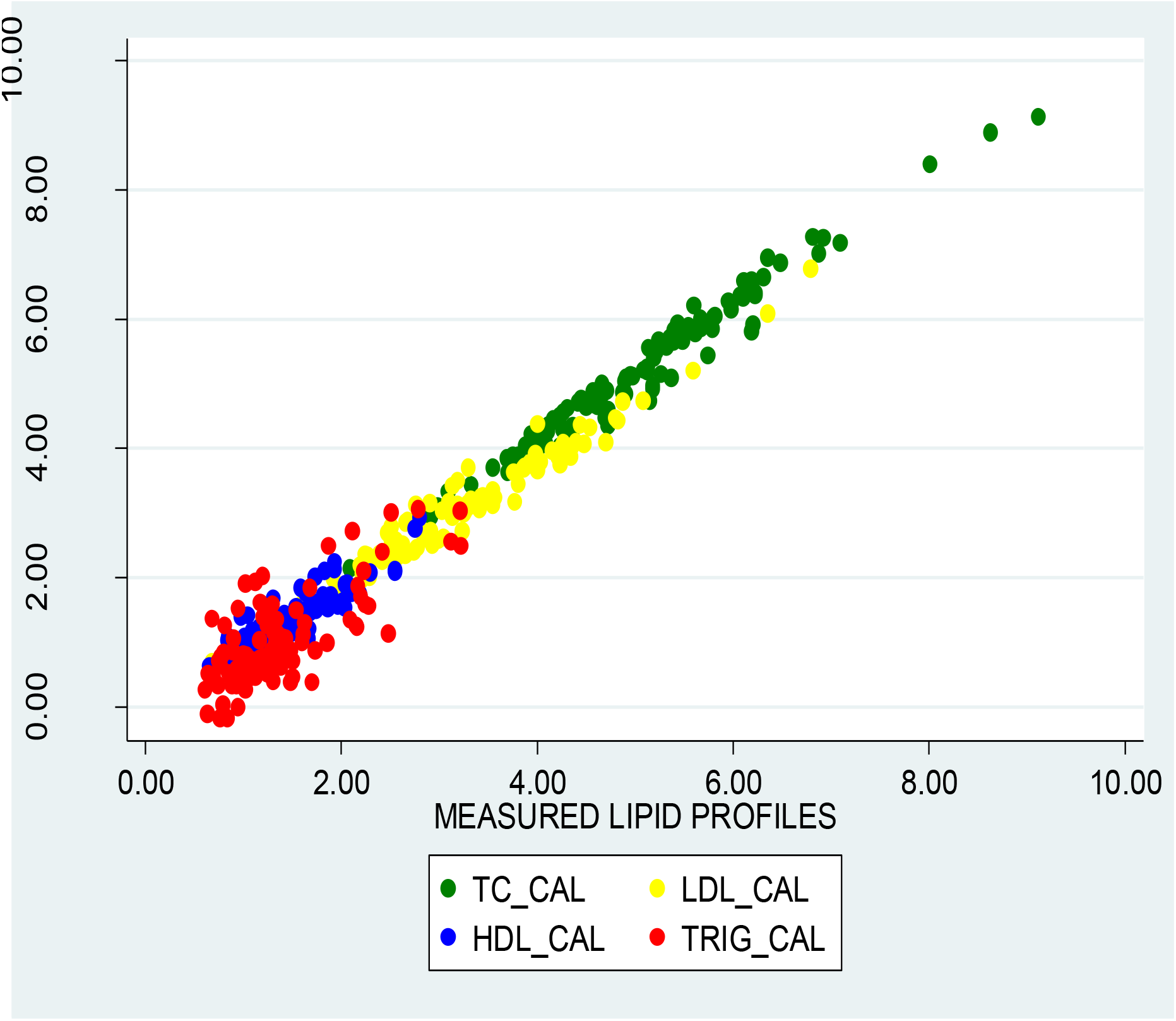
A scatter plot of calculated lipid profile values against measured lipid profile values **Key: LDL-CAL; calculated LDL-C, Trig-CAL; calculated triglycerides, HDL-CAL; calculated HDL-C, TC-M; measured total cholesterol**

### 3.4 ACCURACY OF EXPANDED FRIEDEWALD EQUATION IN CARDIOVASCULAR DISEASE RISK STRATIFICATION

Sensitivities, specificities, likelihood ratios and ability of calculated lipid profile tests to correctly classify subjects according to the NCEP ATP III guidelines are highlighted in table 4. Cut-offs used to classify patients were from NCEP ATP III guidelines. LDL-CAL highest sensitivity was for values < 2.59 mmol/L (optimal risk category) at 90.63% while its lowest sensitivity was for values 4.14 to 4.89 mmol/L (high risk category) at 35.71% while TC-CAL had its highest sensitivity for values <5.17 mmol/L (desirable risk category) at 92.86 and its lowest sensitivity for values 5.17 to 6.18 mmol/L (borderline high risk category) at 77.78%. HDL-CAL highest sensitivity was 73.33% for values <1.03 mmol/L (low risk category) and lowest at 56.41% for values >1.55 mmol/L (high risk category). Highest percentage of correctly classified subjects was for LDL-CAL values > 4.91mmol/L (very high risk category) at 99.03 while lowest was for HDL-CAL values <1.03 mmol/L (low risk category) which was 75.73%. Highest positive likelihood ratio was for LDL-CAL values >1.55 mmol/L (high risk category) at 36.1026 while lowest positive likelihood ratio was for HDL-CAL values <1.03mmol/L (low risk category) at 3.073.

**Table 4:** Sensitivity, specificity, likelihood ratios (LR+ and LR-) and percentage of correctly classified subjects by calculated LDL, TC and HDL across risk categorization concentrations established by NCEP ATP III guidlines.

| **<u>Risk categorization</u>** | **<u>Sensitivity</u>** | **<u>Specificity</u>** | **<u>Correctly Classified</u>** | **<u>LR+</u>** | **<u>LR-</u>** |
| --- | --- | --- | --- | --- | --- |
| **LDL < 2.59; Optimal risk** | 90.63% | 91.55% | 91.26% | 10.7240 | 0.1024 |
| **LDL 2.59-3.34; Above optimal** | 73.53% | 84.06% | 80.58% | 4.6123 | 0.3149 |
| **LDL 3.36-4.11; Borderline high** | 52.63% | 85.71% | 79.61% | 3.6842 | 0.5526 |
| **LDL 4.14-4.89; High** | 35.71% | 97.75% | 89.32% | 15.8928 | 0.6576 |
| **LDL > 4.91; Very High** | 75.00% | 100.00% | 99.03% |  | 0.2500 |
| **TC < 5.17; Desirable** | 92.86% | 91.49% | 92.23% | 10.9107 | 0.0781 |
| **TC 5.17-6.18; Borderline High** | 77.78% | 92.11% | 88.35% | 9.8519 | 0.2413 |
| **TC CAL >= 6.18; High** | 87.50% | 93.10% | 92.23% | 12.6875 | 0.13430 |
| **HDL < 1.03; Low** | 73.33% | 76.14% | 75.73% | 3.0730 | 0.3502 |
| **HDL > 1.55; High** | 56.41% | 98.44% | 82.52% | 36.1026 | 0.4428 |

Key: LDL-M; measured LDL-C, LDL-CAL; calculated LDL-C, Trig-M; measured triglycerides, Trig-CAL; calculated triglycerides, HDL-M; measured HDL-C, HDL-CAL; calculated HDL-C, TC-M; measured total cholesterol, TC-M; measured total cholesterol

## 4.0 DISCUSSION

In this study, total cholesterol and LDL-C presented with calculated means statistically similar to those measured boosting their utility in lipid profile testing. Calculated and measured HDL-C and triglycerides had statistically unequal means which lowers the utility of Friedewald equation in their calculation. The mean value of LDL-CAL was lower than LDL-M despite being statistically similar which agrees with findings by Gupta et al and Vujovic et al ^14,16^. However, variation was demonstrated in a study performed on a Ghanaian population where the mean LDL-CAL was significantly higher than LDL-M ^11^. Iranian population results were as well contradictory to our findings with a higher LDL-CAL than LDL-M ^17^.

TC-CAL and LDL-CAL total errors were below respective NCEP error limits of ±9% and ±12% that continues strengthen utility of the equation. However, HDL-CAL and trig-CAL total errors were above respective NCEP error limits of ±13% and ±15%. Trig-CAL total error level was higher than CLIA set limit of ±25% while HDL-CAL was below its CLIA set limit of ±30% which further derails the equation’s use in triglyceride calculation but allows for calculating HDL-C ^18^. TC-CAL and LDL-CAL were below respective CLIA errors limits of 10% and 30% giving more confidence to calculation of LDL-C and total cholesterol ^18^. Total error between calculated and measured LDL-C were in a similar range compared to those established by Vujovic et al who found this difference to be 6.9% ^16^ and that established by Ephraim et al who showed a mean 12.39 ^11^. Total error differences of calculated LDL-C may be attributed to population differences which has prompted modifications in the equation to best suit respective populations. However, modifications show little variations from Friedewald equation ^19^ which remains vital in populations without modified equations.

In comparison to measured values across NCEP ATP III established concentrations for cardiovascular disease risk stratification, TC-CAL had the best average sensitivity and specificity, followed by LDL-CAL and HDL-CAL. LDL-CAL sensitivities and specificities were similar to those established by Martins et al but inconsistent with Ephraim et al findings that showed lower sensitivities and specificities across LDL-C stratification categories ^11,20^. Atleast 75% persons were correctly classified and calculated values across most stratifications categories yielded high positive likelihood ratios above 9 and low negative likelihood ratios below 0.7 which strengthen the equation’s use. However, LDL-CAL in the range of 2.59 to 3.34 mmol/L, 3.36 to 4.11 mmol/L and HDL-CAL less than 1.03 mmol/L showed positive likelihood ratios of 4 and below which potentially restricts the equation’s use at these concentrations.

Significant strong correlation between calculated and measured values lipid profile tests was observed. This gives Friedewald equation strength in calculation of lipid profiles. However, high Trig-CAL error beyond published error limits potentially out-weigh its correlation strength. LDL-C correlation findings are consistent with findings by Ephraim et al and Krishnaven et al that demonstrated strong positive correlations ^11,21^. Ephraim et al demonstrated a lower calculated LDL-C correlation below 0.9 compared to our findings and those reported by Krishnaven et al ^11,21^. Significant strong positive correlation between measured and calculated lipid profile values among HIV positive and HIV negative persons was seen allowing expanded use of the Friedewald equation. Average correlations of HIV positive persons was statistically similar to that of HIV negative individuals. Findings were concistent with routine practice where an HIV test may not be performed before LDL-C calculation. Scott R. Evans et al also showed a strong positive correlation between calculated LDL-C values and directly quantified LDL-C values among HIV positives which is consistent with our findings ^22^.

Efforts to trace studies that calculated lipid profile tests other than LDL-C using the Friedewald equation were futile giving this study a potential strength of being the first to expand the utility of Friedewald equation. Additionally, to our knowledge, no studies have compared measured LDL-C to calculated LDL-C values among HIV positive and HIV negative persons 50 years and older who are a high risk population for cardiovascular disease. However, the study has limitations of using a low sample size in comparison to related studies on the Friedewald equation and only examining applicability of the equation at triglyceride levels below 4.52mmol/L or 400 mg/dl.

## 5.0 CONCLUSION

The Friedewald equation is a reliable estimator of total cholesterol, LDL-C and HDL-C but not triglycerides among the elderly. The equation yields more accurate total cholesterol results than LDL-C for which it was designed. This utility of Friedewald equation cuts across both HIV positive and HIV negative individuals. However, there is need to further study this utility of the Friedewald equation across different ages, triglyceride concentrations and larger sample sizes

## Data Availability

Data will be availed upon request

## 6.0 DATA AVAILABILITY

Lipids paper.excel document attached.

## 8.0 ACKNOWLEDGEMENT

Authors acknowledge Mildmay Uganda laboratory staff and the Mildmay Uganda scientific advisory and research committee for making this work possible. This research did not receive any specific grant from funding agencies in the public, commercial, or not-for-profit sectors.

## 9.0 AUTHOR CONTRIBUTIONS

**Emunyu Jude**; Concept development, data analysis, manuscript draft and revision

**Semigga Brian**; Concept development and data collection

**Bakyayita Charles**; Supervisory role

**Emmanuel Omony**; Supervisory role

**Namukwaya Brenda:** Manuscript revision

**Kisembo Stephen**; Manuscript revision

**Namubiru Bridget**; Manuscript revision

